# Simple ventilators for emergency use based on Bag-Valve pressing systems: Lessons learned and future steps

**DOI:** 10.1101/2020.04.29.20084749

**Authors:** E. Castro-Camus, J. Ornik, C. Mach, G. G. Hernandez-Cardoso, B. Savalia, J. Taiber, A. Ruiz-Marquez, K. Kesper, S. Konde, C. Sommer, J. Wiener, D. Geisel, F. Hüppe, G. Kräling, J. Nguyen, T. Wiesmann, B. Beutel, M. Koch

## Abstract

As part of a plethora of global efforts to minimize the negative effects of the SARS-CoV2 (COVID-19) pandemic, we developed two different mechanisms that, after further development, could potentially be of use in the future in order to increase the capacity of ventilators with low-cost devices based on single-use-bag-valve mask systems. We describe the concept behind the devices and report a characterization of them. Finally, we make a description of the solved and unsolved challenges and propose a series of measures in order to better cope with future contingencies.

## Introduction

The international health contingence caused by the rapid propagation of the SARS-CoV2 (COVID-19) has triggered an unprecedented type of collaborative and distributed development worldwide. From professional scientists, physicians and engineers in universities, research institutions and companies to individuals with home-based electronics and mechanics building capabilities have started intensive brainstorming, prototyping, re-engineering and testing of potential solutions to multiple aspects of the pandemic.

Taking into account the experience gained in China and Italy, two of the earliest and most affected countries by the pandemic so far, it is clear that a key piece of equipment required at hospitals are mechanical ventilators and that the number of available ventilators is insufficient with regard to the potential numbers needed in most countries worldwide.^1^ On 13.03.2020, we decided to start an emergency project to develop and evaluate the potential of a platform for low-cost, easy-to-build ventilators to cope with the potential shortage of ventilators worldwide. During the following days, an interdisciplinary group was formed including people with backgrounds in Physics, Engineering, Informatics, Medicine, etc. to shape up ideas. Quickly two approaches were chosen, prototypes started to be built, tested and optimized for performance and reliability. In this contribution we present two ventilator prototypes of the most basic approach which are based on the use of a Bag-Valve manual ventilator.

The design framework shown here consists of the aspects of low cost design, easy modification of widespread standard components, or at least easy to manufacture, and basic ventilator capacity. In addition, we enumerate challenges pending to be solved for this concept to be converted into a real solution. It is worth mentioning that while we have tried to include as many of the options and to meet characteristics similar to a professional mechanical ventilator, our objective was to build a simplified, easy- and economical-to-build apparatus that, when fully developed, could potentially be of benefit to patients in need for mechanical ventilation in a circumstance in which no other option can be offered. We also want to make a clear statement that the devices presented here:

1. Have not been approved for use by any health regulatory agency or homologous institution.
2. Have not been tested on humans or other animals, and that all tests have been carried out with mechanical and electronic equipment only. No clinical or preclinical trials have been conducted or approved up to the date of this publication.
3. We strongly discourage any attempt to use the devices presented here on humans. Its replication should only be for the purpose of further investigation and development.

Manually pumped ventilators have been used in the past to cope with epidemic-related patient surges.^2^ It is also good noticing that pushing mechanisms for a bag-valve ventilator is an approach that has been pursued by various groups around the world both in the past, mostly as an academic exercise.^3-5^ Yet, in recent days many groups around the world have taken this concept as a possibility to cope with the current crisis^6-9^. It is worth mentioning that our designs might have similarities and face similar deficiencies to other bag-valve pushing mechanisms. For some of those developments detailed construction schematics and other detailed information have been made available to the public.

## Design considerations

While the use of mechanical ventilation is a clinical practice that is normally only used on patients that require it as a life-sustaining measure, inappropriate ventilation of a patient could result in damage to the lungs that would also be life-threatening.^10-12^ The breathing process might seem a relatively simple one at first glance. To first approximation it is not very different to inflating and deflating a balloon, however, that simplistic view is what physicists would call a spherical-cow-approximation. There are a number of subtleties that make respiration more complex. In the following, we provide a brief description of the breathing process based on^13^. Under normal circumstances, inhalation is driven by the unconscious motion of a series of muscles, mainly the diaphragm as the key muscle together with different other muscle groups of the chest wall, that increase the volume of the thoracic cavity. This leads to a decrease of the pressure in the lungs which, at its time, results in the flow of fresh air into the lungs through the airway. The subsequent relaxation of said muscles increases the pressure in the lungs that produces the exhalation. Therefore inhalation is usually produced by “negative” thoracic pressure, while exhalation is produced by “positive” thoracic pressure. When mechanical ventilation is applied, usually through an endotracheal tube, the inhalation is no longer driven by negative pressure produced by the patient, but by positive pressure provided by the ventilator that “forces” air into lungs, this is a complete game-changer in many senses. For instance, natural “negative” pressure inspiration can not over inflate the lungs and its internal ramified structures, the alveoli, which can potentially be damaged by overpressure when “positive” pressure is applied. This could lead to damage of the lung, which is usually already compromised by the underlying condition, such as pneumonia caused by COVID-19. Therefore, the volume per inspiration (tidal volume) and the appropriate pressures across the breathing cycle must be limited.

Unlike healthy subjects, who maintain a small volume of air in the lungs once the exhalation is over, critical patients in some cases produce a full exhalation, this causes the fluids that are in the lung to completely fill all the cavities, which means that it is more difficult to inflate the lung again in the following breath cycle, and making it more prone to injury by the mechanical ventilation, therefore, a minimal positive pressure (Positive End-Expiratory Pressure; PEEP) should be maintained in the lung at the end of the expiration in order to prevent the collapse of the lungs. It is worth mentioning that bag-valve resuscitators are single-use-products, thus, for each new patient, a new resuscitator is needed. Yet, hygienic standards are easy to follow, as the two mechanism prototypes themselves are not part of the pneumatic system itself. In addition, a connecting hose between the bag-valve system and the mask or endotracheal tube will be needed, which is normally not used in manual resuscitation. The total volume of that hose has to be significantly smaller than the tidal volume pumped in order to ensure appropriate exhaustion of the exhaled CO_2_ to the atmosphere.

In addition to the purely pneumatic considerations, there are other aspects to be taken into account. For instance, forcing air into the lungs of a patient at a rate determined by the ventilator instead of the natural rhythm determined by the patient is extremely uncomfortable and requires deep sedation, which might complicate other aspects of the progress evaluation and treatment of the patient. Hence, a feedback mechanism, usually by pressure or flow, is desirable in order to trigger the breath cycle of the ventilator and thus improve the patient-ventilator synchrony. Furthermore the gas used for ventilation should be enriched in oxygen content (FiO_2_) since the lung capacity of the patient is diminished, and ideally the gas mixture should be humidified and warmed-up since the endotracheal tube bypasses the upper part of the airway (nose & pharynx), which usually performs these two functions.

During the four weeks that the prototyping process took in our group, we tried to incorporate as many of the desirable characteristics as possible to our prototypes, however, each degree of sophistication increases the cost of the device, the fabrication time and most importantly the potential failure points. In the face of the contingence careful reliability tests can not be thoroughly performed for appropriately long periods. In Table 1 the typical setting parameters for a mechanical ventilator are shown^14^

**Table 1.** Setting parameters for a mechanical ventilator^14^. * Breaths per minute. ** Acute Respiratory Distress Syndrome.

| Parameter | Value |
| --- | --- |
| Tidal volume | 6-8 ml/kg PBW |
| Rate | 2-16 BPM*, for ARDS** up to 35BPM* |
| PEEP | 5-10 cm H<sub>2</sub>O |
| FiO<sub>2</sub> | Adjusted to obtain SpO<sub>2</sub> >90% |
| Inspiratory flow | 40-60 l/min |
| Insp:exp ratio | 1:1 - 1:3 |
| PSV | 5-10 cm H<sub>2</sub>O |
| Insp. pressure | 12-25 cm H<sub>2</sub>O (typ) |
| Trigger pressure | -2 to -1 cm H<sub>2</sub>O |
| Trigger flow | 2 l/min |

## Results

### Prototype 1 (Contingence Life-support Air Pressure Emergency Respirator; CLAPER)

The first approach we demonstrate here was the construction of a bag-valve pusher that would be able to operate in continuous mode without the requirement of digital control, at least for a first simplified version, even if it did not meet all the ideal requirements. The system consists of a motor that drives a small gear, which at its time drives two larger gears coupled to each other in order for them to move synchronously but in opposite directions. The two gears have an additional eccentric axis that is used to move a coupling rod each, which moves an oscillatory arm which is fixed at one end to the baseplate of the mechanical system. A convex piece is attached to the other end of the arm which is used to press the bag-valve at regular intervals. The position of the fixed point of the arm can be varied and this provides the user with control over the tidal volume (see the methods section). The frequency can be varied as well by changing the voltage supplied to the motor. The ensemble is shown in Fig. 1. However, the inclusion of a microcontroller opens the possibility of controlling not only the breathing frequency and the volume pumped, but the inspiration/expiration time ratio, pressure limits, and pressure/flow triggers.

**Figure 1.**
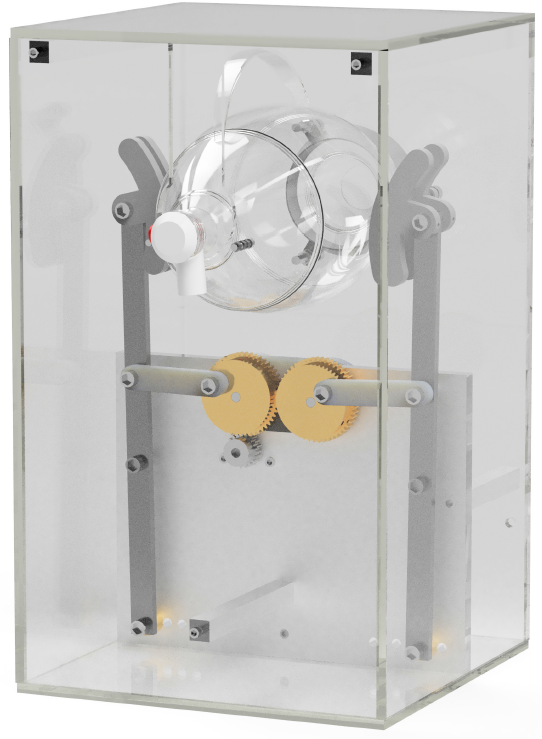
Three-dimensional render of the prototype of CLAPER.

Measurements of pressure and flow were performed on the CLAPER device in order to quantify its performance and decide if it is within the expected parameters for a mechanical ventilator as described in the Methods section. The measurement results are shown in Fig. 2. The tests were performed using the different positions of the pivot in order to have some control over the tidal volume pumped which is a parameter that needs to be controlled for its use. Fig. 2(a) shows the time evolution of the pressure along 4 breathing cycles for various positions of the pivot. The points in Fig. 2(b) show the peak pressure achieved for the different pivot positions, showing that peak pressures between 8.9 and 25.9 cmH_2_O were achieved. Fig. 2(c) shows the evolution of the flow for the breathing cycle and the peak inspiratory and expiratory flows are shown in panel (d). From those measurements it is possible to obtain the evolution of the volume pumped, which is shown in Fig. 2(e). The peak values of that evolution, shown in Fig. 2(f) are the tidal volumes that were achieved for the different positions of the pivot, in this same plot we included gray boxes that depict the tidal volume ranges for patients of different predicted-body-weights which are labeled for each box as a reference. As observed in the plot, by varying the positions it is possible to modulate the tidal volume stepwise for patients from about 30 kg to about 100 kg.

**Figure 2.**
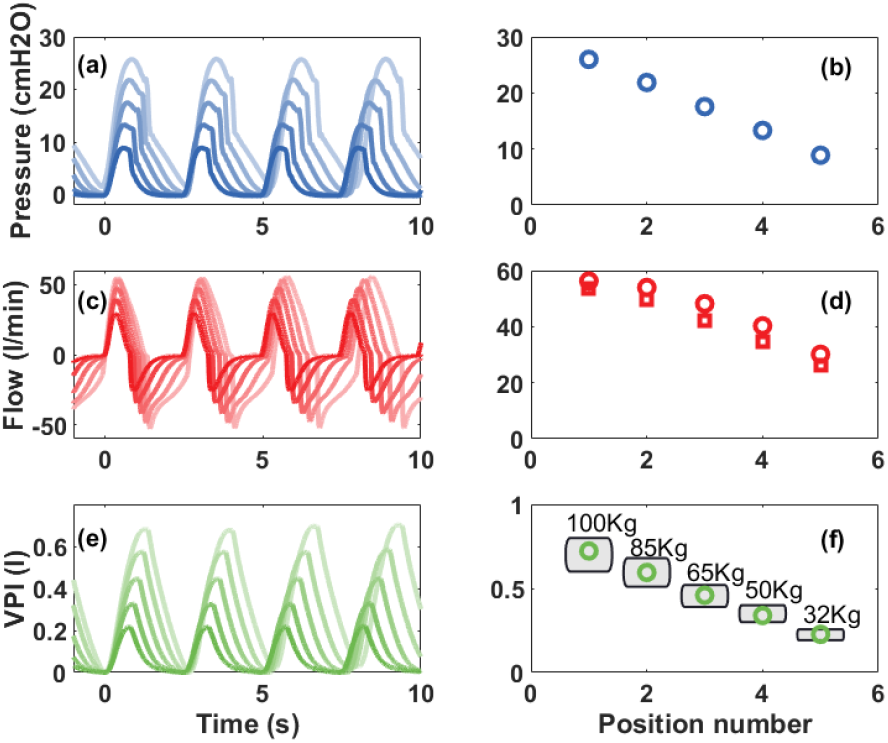
(a) Pressure as a function of time for pivot positions 1 (lighter color) to 5 (darker color). (b) Peak pressure from plots in (a) as function of pivot position. (c) Flow generated as a function of time for pivot positions 1 (lighter color) to 5 (darker color). (d) Peak inspiratory (circles) and expiratory flow (squares) as function of pivot position. (e) Volume pumped as a function of time for pivot positions 1 (lighter color) to 5 (darker color). (f) Peak volume pumped for the various pivot positions, the gray boxes show the recommended tidal volumes (6 to 8 ml/kg) for various predicted body weights (ideal weights) calculated from the parameters in Table 1.

An additional test that was carried out is to measure the maximum obtainable pressure from the system. It is important to mention that this is related to the motor used. In particular we drive our prototype with a ~60 RPM that can deliver a maximum torque of 180 N cm. If a stronger motor is used, that will provide a higher maximum pressure, and vice versa. The measurement was carried out by blocking the outlet of the system, which gives the same effect as an infinitely stiff lung, the maximum pressure we achieved under the given conditions was 44.8 cm H_2_O, which is comparable to commercially available ventilator settings.

### Prototype 2 (experimental Actuated Resuscitator Machine; eARM)

The second approach is based on a simpler mechanical design. The mechanism consists of two arms that squeeze the bag-valve resuscitator, which is placed between them. The motion of the arms is driven by a string that is attached to a string role driven by a motor. Once the resuscitator is squeezed, the direction in which the motor is rotating has to be reversed. In the simplest case, this can be achieved using a microswitch, which marks the end position and relays, which switch the polarity of the voltage applied to a DC motor. A more advanced approach, which we present here, includes using a microcontroller and an H-bridge to invert the motion of a DC motor. Placing two microswitches at the two ends of the arms’ travel, in the initial (open) and final positions (closed), enables positional feedback to a microcontroller. By changing the position of the microswitch, which indicates the final (closed) state, the pressure and tidal volume can be tuned. The use of a microcontroller enables the control and monitoring of the breathing rate, inspiration to expiration ratio, and opens the possibility to implement complex feedback-based controls (e.g. PEEP control, triggered inspiration). A render of the prototype is shown in Fig. 3.

**Figure 3.**
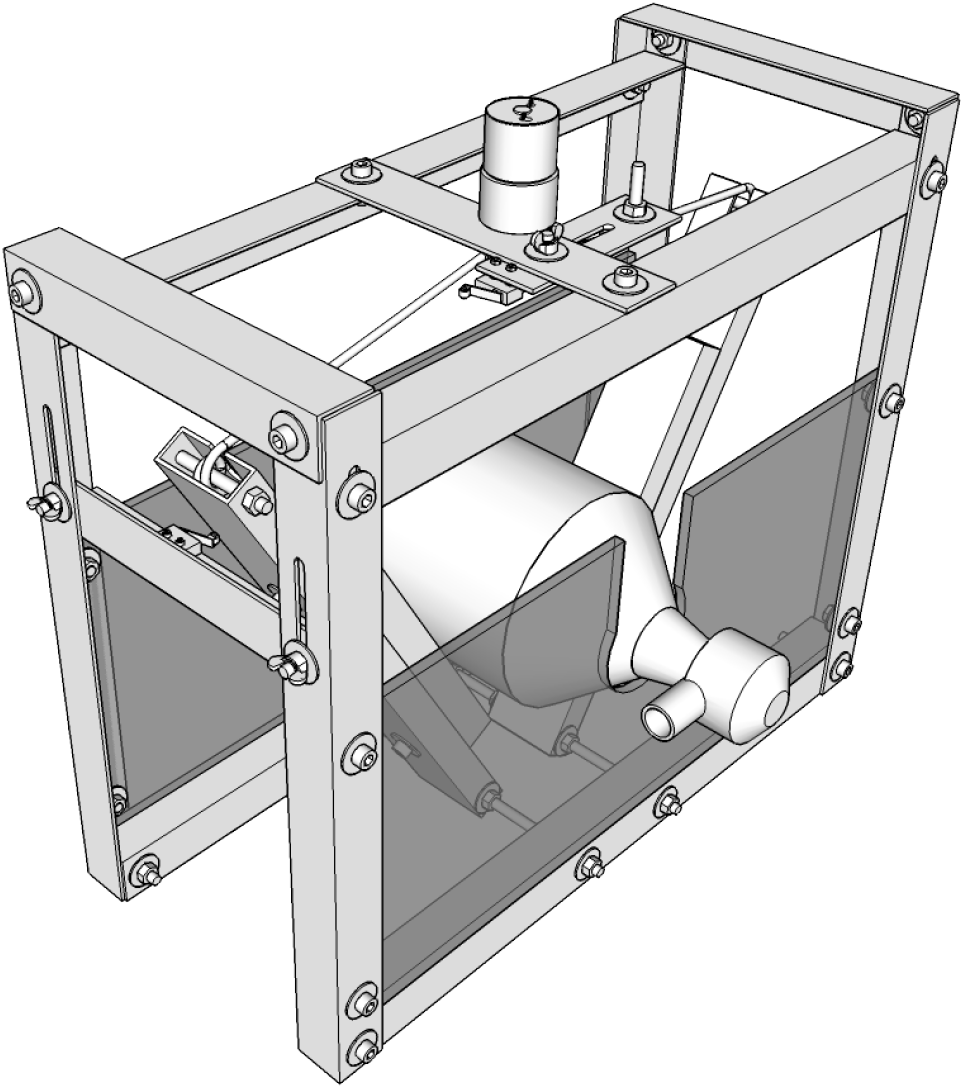
Three-dimensional render of the eARM prototype.

Measurements of pressure and flow were also performed on the eARM using the same equipment as for CLAPER as describen in the Methods section. An additional tube of approximately 2 m length and 1 cm in diameter was used for connecting the test lung to the bag-valve resuscitator. An additional valve from another bag-valve resuscitator was placed between the tube and the test lung, in order to prevent “exhaled” air from residing in the tube and being “inhaled” with the next inspiration. The tests measurements were performed at five different positions of the microswitch indicating the final (closed) position in order to control the tidal volume. The breathing rate was set to 20 BPM and the inspiration to expiration ratio to 1:1. The measurement results are shown in Fig. 4. In the left part of Fig. 4 the time evaluation of pressure, flow, and volume are shown over 4 breathing cycles for the five positions of the microswitch. The average of the peak pressure, peak volume, and peak inspiratory and expiratory flow for each position of the microswitch are shown in the right part of Fig. 4. Since both prototypes rely on a bag valve resuscitator, a similar outcome of the test measurements was expected and also achieved. As a microcontroller is already used for this prototype, an additional pressure sensor was installed. This way we were able to show the potential of such a device to also control PEEP as well as potential for triggered inspiration (Augmented spontaneous breathing [ASB] mode). The test measurements with the implemented pressure control are shown in Fig. 5.

**Figure 4.**
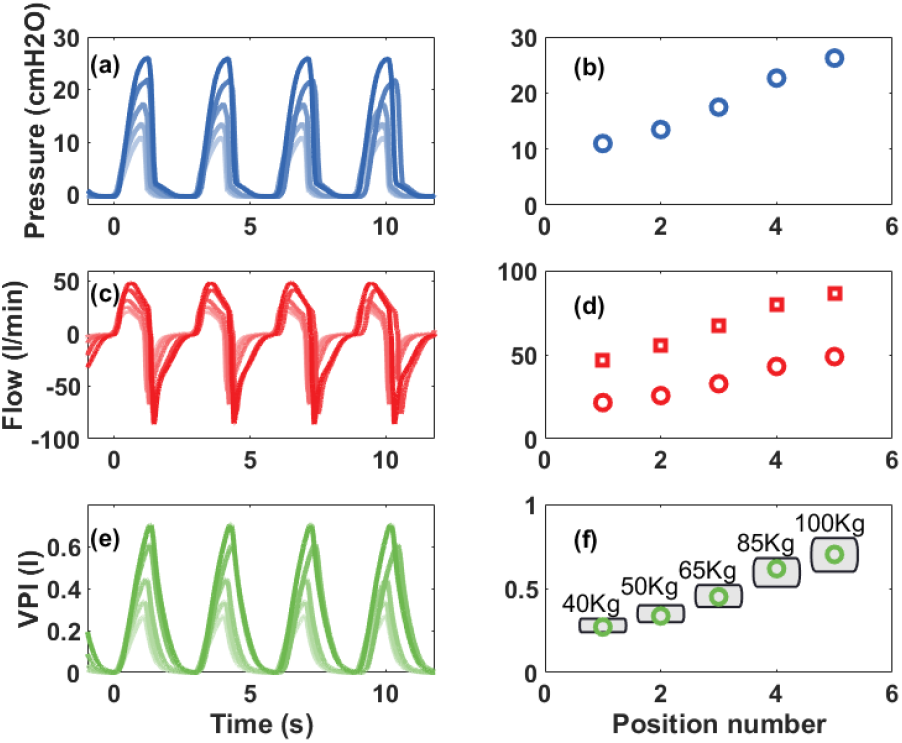
eARM measurement results. (a) Pressure as a function of time for microswitch positions 1 (lighter color) to 5 (darker color). (b) Peak pressure from plots in (a) as function of microswitch position. (c) Flow generated as a function of time for microswitch positions 1 (lighter color) to 5 (darker color). (d) Peak inspiratory (circles) and expiratory flow (squares) as function of microswitch position. (e) Volume pumped as a function of time for microswitch positions 1 (lighter color) to 5 (darker color). (f) Peak volume pumped for the various microswitch positions, the gray boxes show the recommended tidal volumes (6 to 8 ml/Kg) for various predicted body weights (ideal weights).

**Figure 5.**
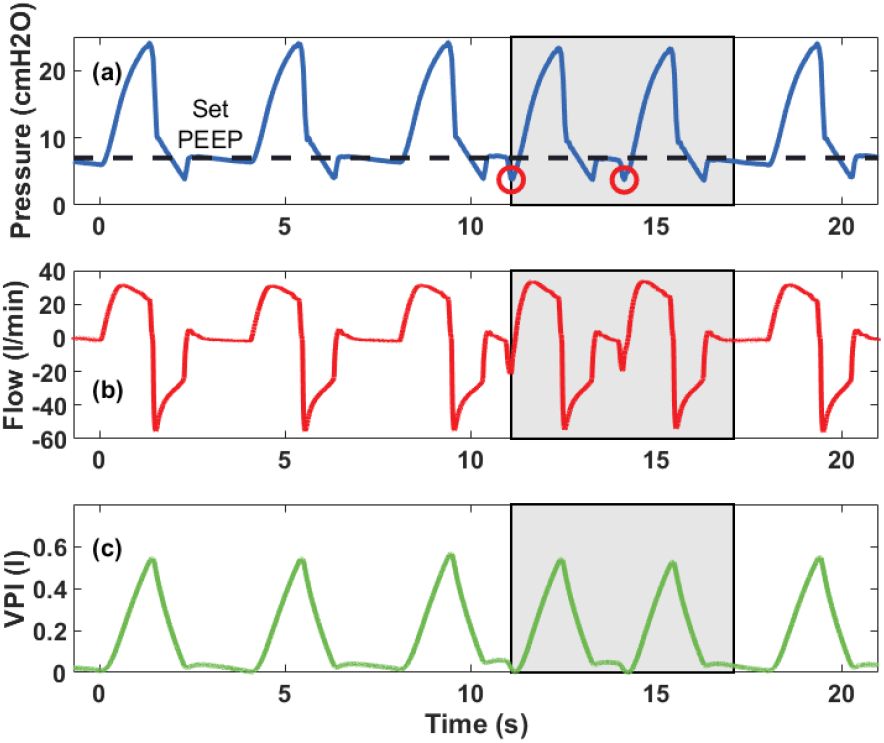
Measurements performed in order to demonstrate the additional features that can be implemented relatively easily with a digital control and a pressure sensor. Respiration rate was set to 15 BPM with inspiration to expiration ratio set to 1:2. The system was also set to trigger a breathing cycle in case of a potential inspiration (marked in gray) and to maintain PEEP, which was done with an additional mechanical PEEP valve and pressure dependent motor control. (a) Pressure obtained along several respiratory cycles. An inspiration was triggered when the pressure dropped by 2 cm H_2_O (red circles) with respect to PEEP. This drop in pressure is the signal that corresponds to a patient attempting to breath in. PEEP was set to 7 cm H_2_O (dashed line). (b) Flow as a function of time. (c) Volume pumped as a function of time.

**Figure 6.**
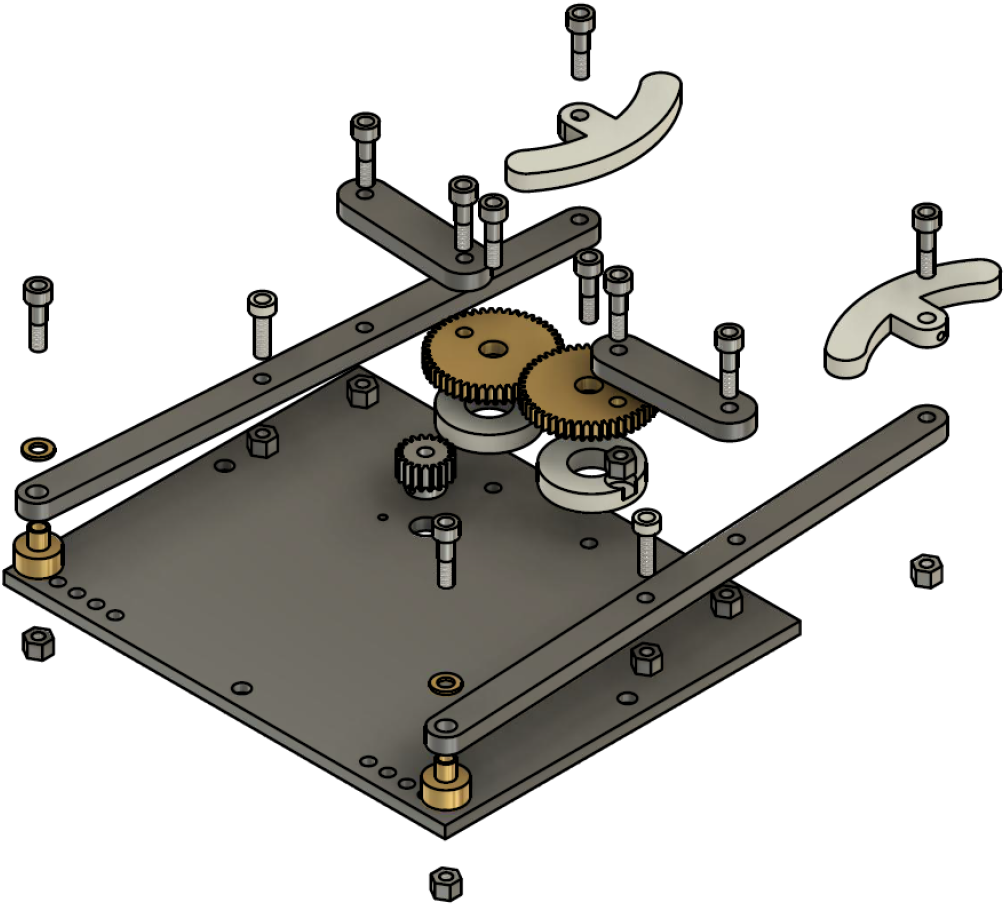
Schematic representation of the assemble. The design will require adjustments.

## Discussion and Conclusions

The performance shown by the two prototypes for the “core” characteristics are comparable to professional ventilators, however there are also some characteristics that are not implemented such as special sequences that help physicians evaluate the state of the patient. The volumes and pressures achieved during the tests are consistent with the clinical requirements, and both systems permit controlling the volumes and breathing rates within the ranges normally used in intensive care units, of course there are still challenges to be overcome in future research:

1. **Reliability of the mechanisms, software, etc.:** Since these were fast track developments, there has not been enough time to undergo appropriate long term reliability tests on these pieces of equipment. Although we did run tests for several days on both prototypes, appropriate tests for weeks or months under realistic operation conditions are still needed and will require a significant amount of time, incompatible with the urgent requirement of ventilators to deal with the ongoing international crisis.
2. **Mass production with appropriate standards:** Given that the concept of the devices presented here with the necessary adjustments for industrial production are expected to become medical equipment, the fabrication has to be performed in appropriate industrial environments with the necessary manufacture equipment and by trained personnel. In addition appropriate quality control has to be implemented. Once again, it is not clear how this could be done in a timely manner to face the COVID-19 pandemic.
3. **Regulatory approval or exemption:** Being medical equipment, the use of the devices presented here, or variations around them, requires appropriate approval by the health regulatory agencies of the different countries that intend to use them. Such a process is, under normal circumstances, time consuming, involves a significant amount of bureaucratic paperwork and financial investment, not to mention extensive preclinical and clinical trials. While some countries, such as the USA have issued a declaration of “liability immunity for activities related to medical countermeasures against COVID-19”,^15^ most countries have not taken similar measures, therefore, the use of these devices depends either on the creation of similar policies of general application, or the expedited approval of specific devices.
4. **Additional features:** In addition to the central characteristics such as volumes, pressures, etc. required from a professional mechanical ventilator, there are a number of other characteristics that need to be met. For instance, the system has to be enclosed in an easy-to-clean fluid spill resistant casing, it requires battery support for the case of a power cut or for patient transportation, among other characteristics. The possibility to provide real-time pressure, flow and volume plots on a screen is also a characteristic that would be desirable as well as the possibility to program other inhalation/exhalation sequences that are of use for clinicians as diagnostic tools.

In conclusion, we believe that our two proof-of-concept approaches to re-engineer and upscale simple low cost manual bag-valve-systems to a low-cost ventilator were successful and provide what promises, after further development, be of benefit to patients in need of mechanical ventilation in the absence of other options. However, a number of questions remain open and this implies the need of reliability tests, clinical trials, as well as fulfilling regulatory and legal formalities. Furthermore, significant commitment from companies that can manufacture them, or even military industry, will be needed if these concepts are to be converted into real solutions within a reasonable timescale. An important aspect is that, perhaps for the first time a global collection of efforts appeared spontaneously in a very short period, and not only to address the mechanical ventilator availability problem, but many other aspects, yet most of these efforts were disorganized and the convergence of the emergency-development, emergency-manufacture and emergency-legal/regulatory-clearance do not seem to be converging at appropriate speeds to deal with the problem. These aspects will need to be addressed in order to better respond to future crises.

## Methods

### Pressure and volume measurements

The measurements were performed by inflating a calibrated MAQUET test lung 190 for a maximum tidal volume of 11, all the tests were conducted using normal atmospheric air at room temperature and pressure. A calibrated digital flow and pressure measurement system was used for the characterization.

### CLAPER prototype

Here we include the schematics of the most important mechanical parts of the design:

1. The baseplate in Fig. 7.
2. The connecting rod in Fig. 8.
3. The arm rod in Fig. 9.
4. The bag pusher in Fig. 10.
5. The gears in Fig. 11.
6. The teflon bearing for the gears in Fig. 12.
7. The bush bearing for the long arms in Fig. 13.

**Figure 7.**
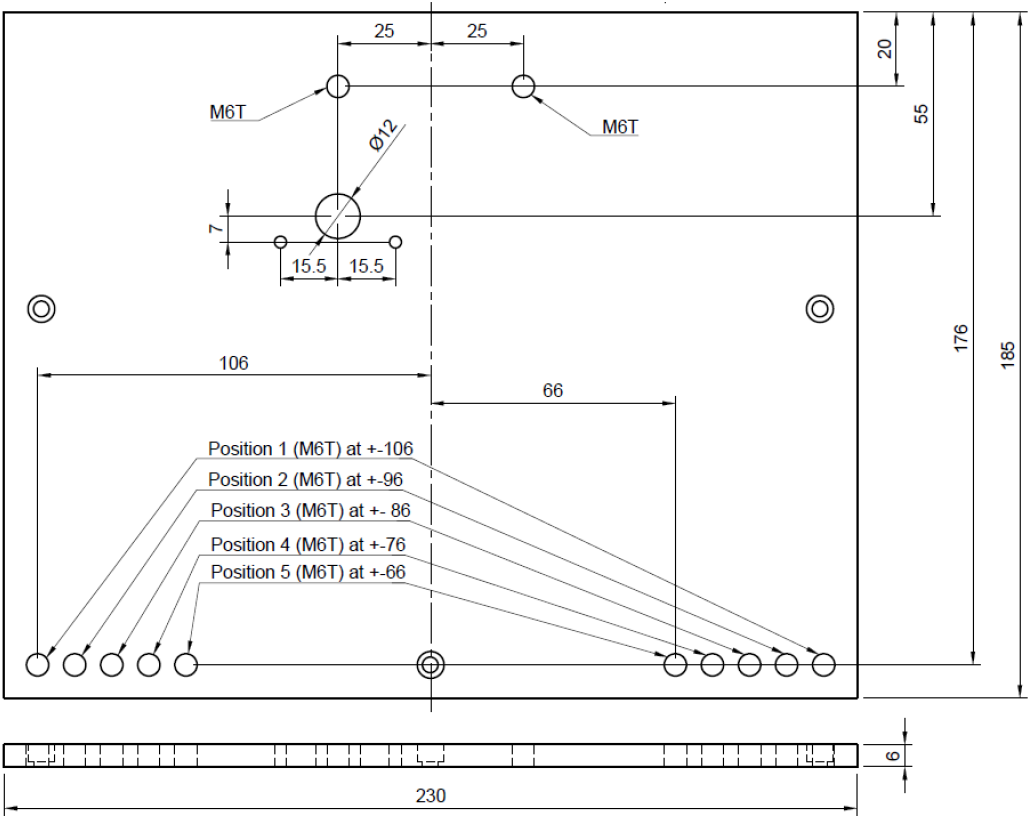
Schematic representation of the baseplate. Sizes shown are just for reference and have not been optimized. The design will require adjustments.

**Figure 8.**
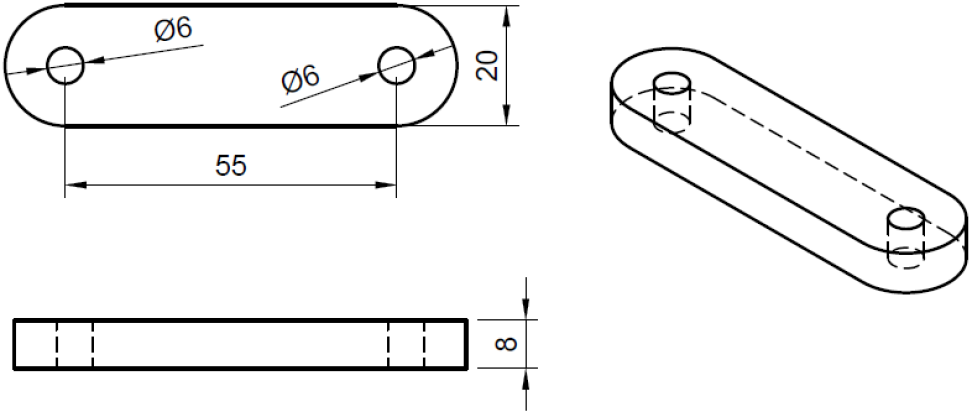
Schematic representation of the connecting rod. Sizes shown are just for reference and have not been optimized. The design will require adjustments.

**Figure 9.**
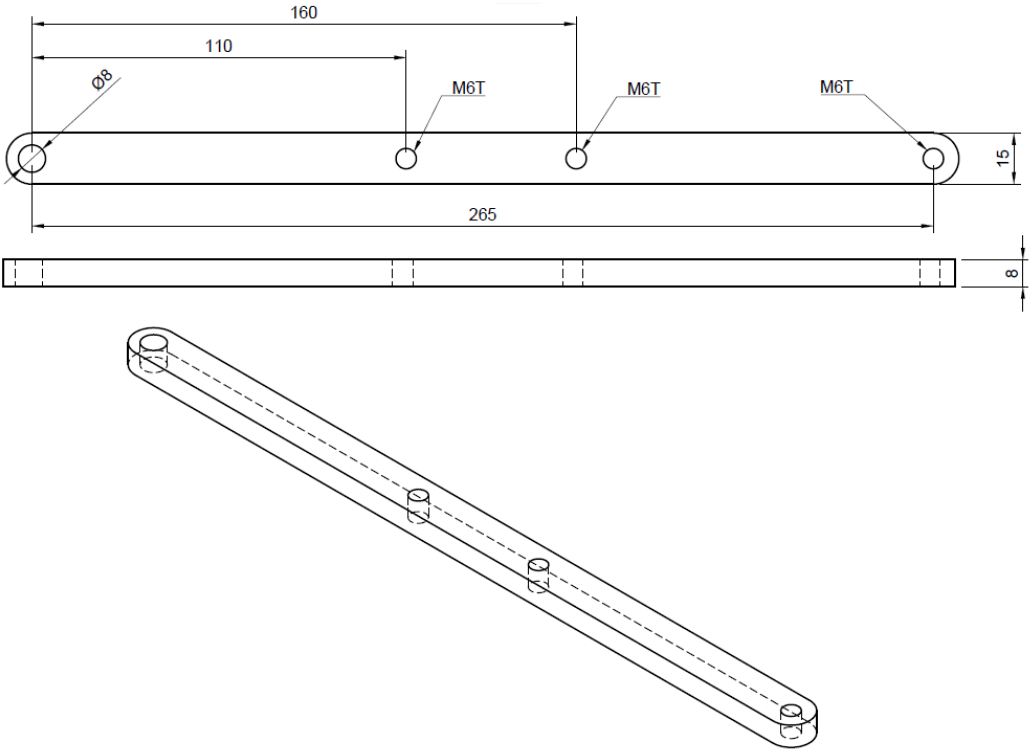
Schematic representation of the arm rod. Sizes shown are just for reference and have not been optimized. The design will require adjustments.

**Figure 10.**
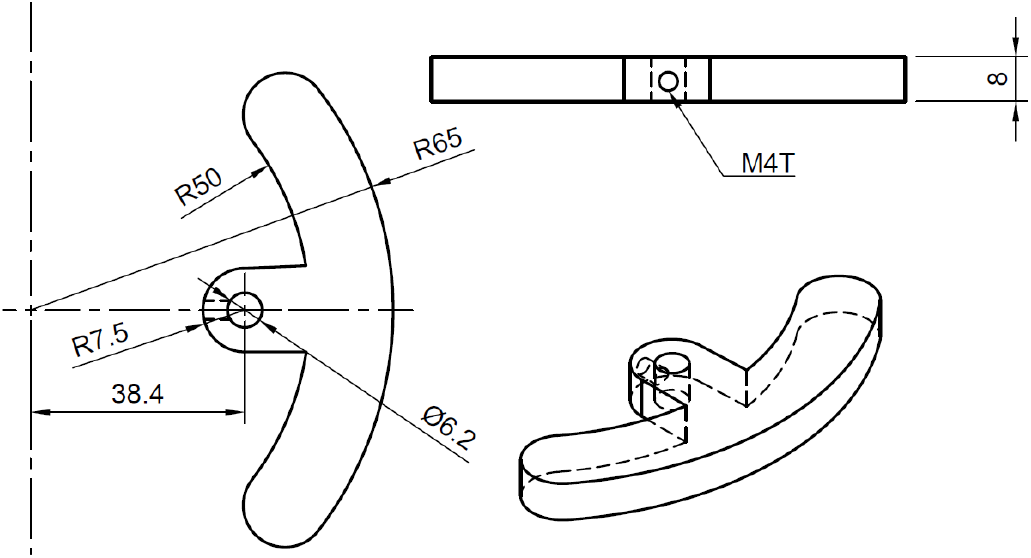
Schematic representation of the bag-pusher. Sizes shown are just for reference and have not been optimized. The design will require adjustments.

**Figure 11.**
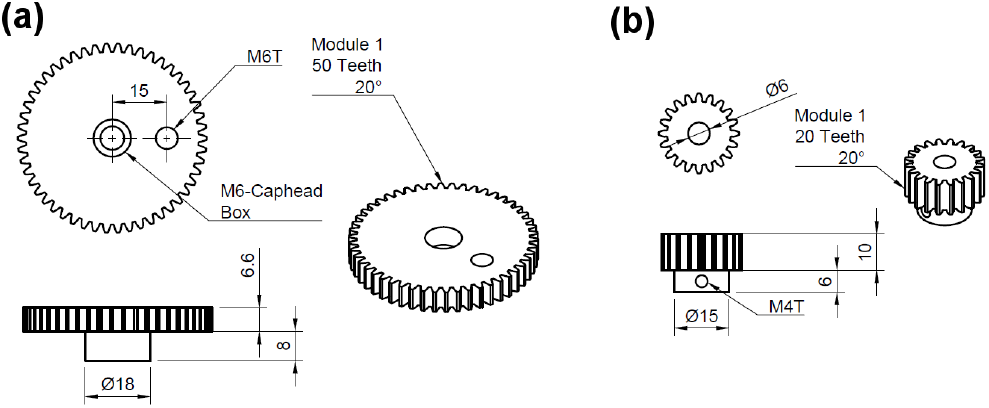
Schematic representation of the gears used. Sizes shown are just for reference and have not been optimized. The design will require adjustments.

**Figure 12.**
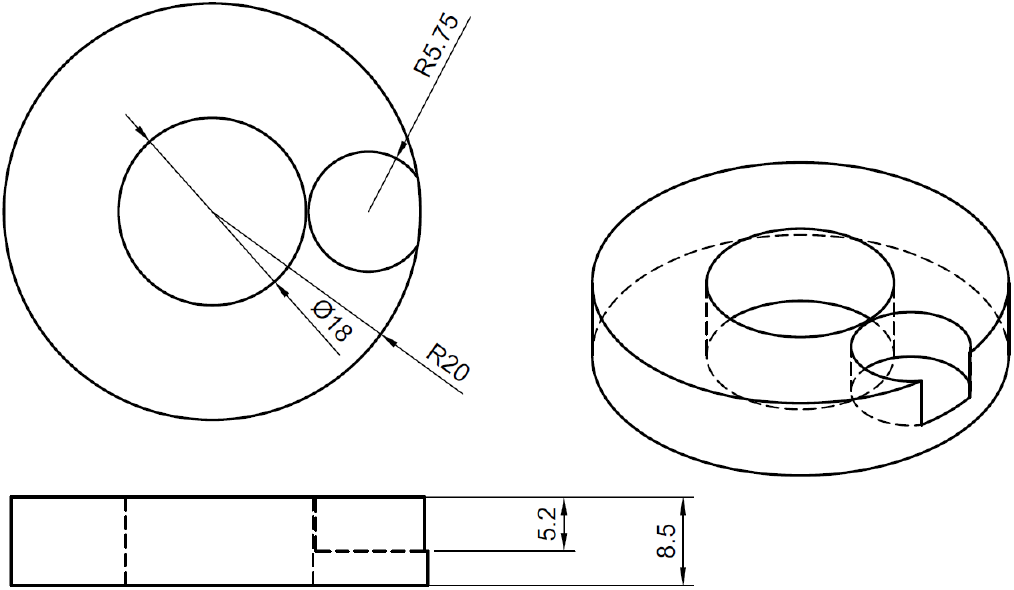
Schematic representation of the teflon-bearing used under the 50-teeth gears. Sizes shown are just for reference and have not been optimized. The design will require adjustments.

**Figure 13.**
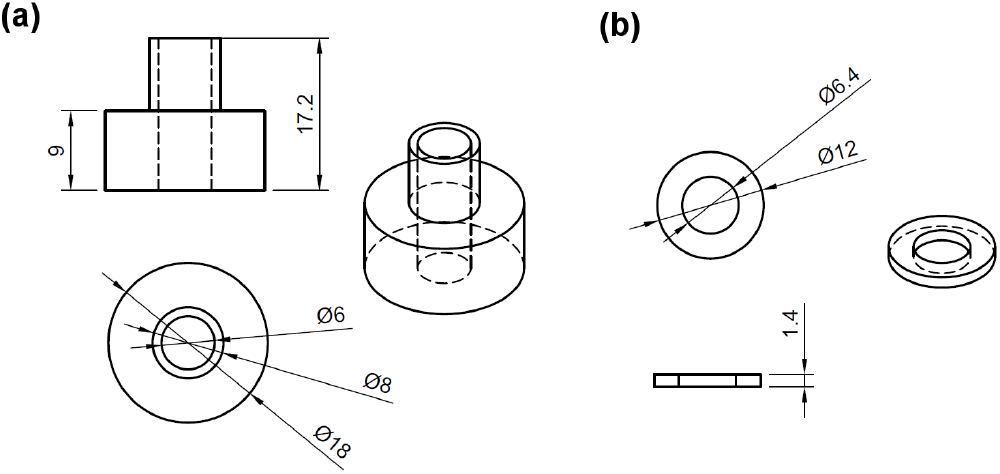
Schematic representation of the bush-bearing used at the long arm pivot position (a) is the main body of the bearing, (b) is a washer that maintains the bar in place. Sizes shown are just for reference and have not been optimized. The design will require adjustments.

The system driven by a DC motor ~60 RPM in the absence of load with a maximum torque of 180 N m. which was coupled directly to the small gear. The mechanical assemble can be seen in Fig. 1 and a schematic showing the details of the assemble process in shown in Fig. 6. For motion axes stainless-steel M6 cap-head screws were used with non-threaded sections. They were fixed on the M6T threads made on the base-plate, gears, or long bars with an additional counter-nut on the opposite side. All parts were machined in aluminium, except for, the bush-bearings and washers (Fig. 13), which were made from brass, the 50-teeth gears, which were made from brass commercial gears, and the 20-teeth gear which was made from a commercial stainless-steel gear. Nylon screws were used at the M6T threads on the long arm 110 mm from the pivot point in order to give support and fine alignment control, also a counter-nut was used to fix the adjustment. A commercial PEEP-valve was fitted to the exhaust of the Bag-Valve in order to have control on the end-exhalation-pressure.

### eARM prototype

Schematics of Prototype 2 (eARM) which is shown in Fig. 14.

**Figure 14.**
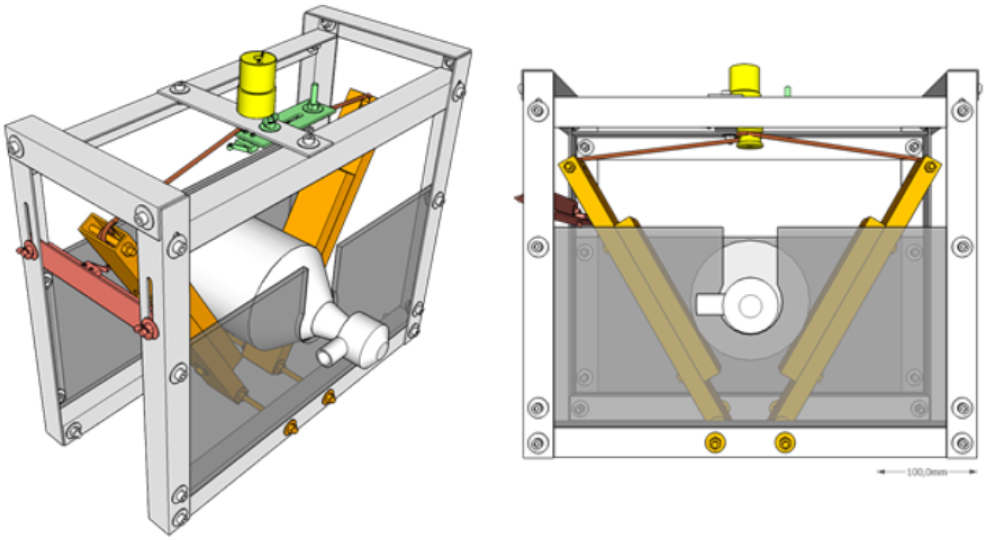
Three-dimensional render of the eARM prototype diagonally and from the front. Yellow: Motor with string roll, Orange: Squeezing arms, Green: Upper endswitch (volume and pressure control), Red: Lower endswitch (Timing control). The design will require adjustments.

Design considerations:

1. A flexible and strong string is required (e.g. braided nylon cord), which should be able to carry enough weight, depending on the pressure which is to be achieved.
2. A spiral form in the string roll (see Figure 15) reduces jitter from string rolling up badly. Choosing a sufficient r2, in a way that squeezing is achieved in only a few turns, also helps avoiding jitter.
3. The string roll should enable easy installation/attachment of the string (e.g. twice through a hole in the string role).
4. An attachment on the arms (b2) helps pressing the bag-valve resuscitator while leaving space for the string roll (g > 2r2)
5. To avoid potential damages of the bag-valve resuscitator at the maximum squeezing, an additional space between the arms (dg) should be left.
6. Movable microswitches make setting up the device easier. The tunable position of the upper microswitch is essential for a manual volume and pressure control.
7. Two microswitches are needed to monitor and set respiration rate and inspiration to expiration ratio. Also if 1:1 ratio is the goal, keep in mind that opening and closing times differ.
8. Since bag-valve resuscitators are not designed for continuous long-term use, their characteristics and material can deteriorate with time. Regular rechecking and if necessary replacement are advised. Therefore, a design enabling relatively easy and quick replacement of it should be realized.
9. In Fig. 16, the arm the length (l), spacing between the two rotational axis (d), bag-valve resuscitator position (h), string roll radius (r) are shown. Accordingly a motor with appropriate characteristics (torque, rpm) have to be chosen in a way, that the squeezing process runs smoothly with the desired pace while reaching the desired pressure and volume.
10. By increasing l, the motor’s leverage also increases, if h stays unchanged. This means that a motor with less torque could perform the squeezing. However, in this case the end points of the arms have to travel a greater distance from released to squeezed state, meaning that the motor has to rotate faster at the same r2 of the string role to maintain the same rate of squeezing (BPM). By increasing the r2, the speed of the motor could be kept unchanged, however, this would cancel out the leverage gained by increasing l. Nevertheless, these relations allow for some tunability of the design according to the needs.
11. The systems requires active control from a microcontroller and we propose the connection schematics shown in Fig. 17. The microcontroller used was the ArduinoUNO platform and the code can be found in the supplementary information.

**Figure 15.**
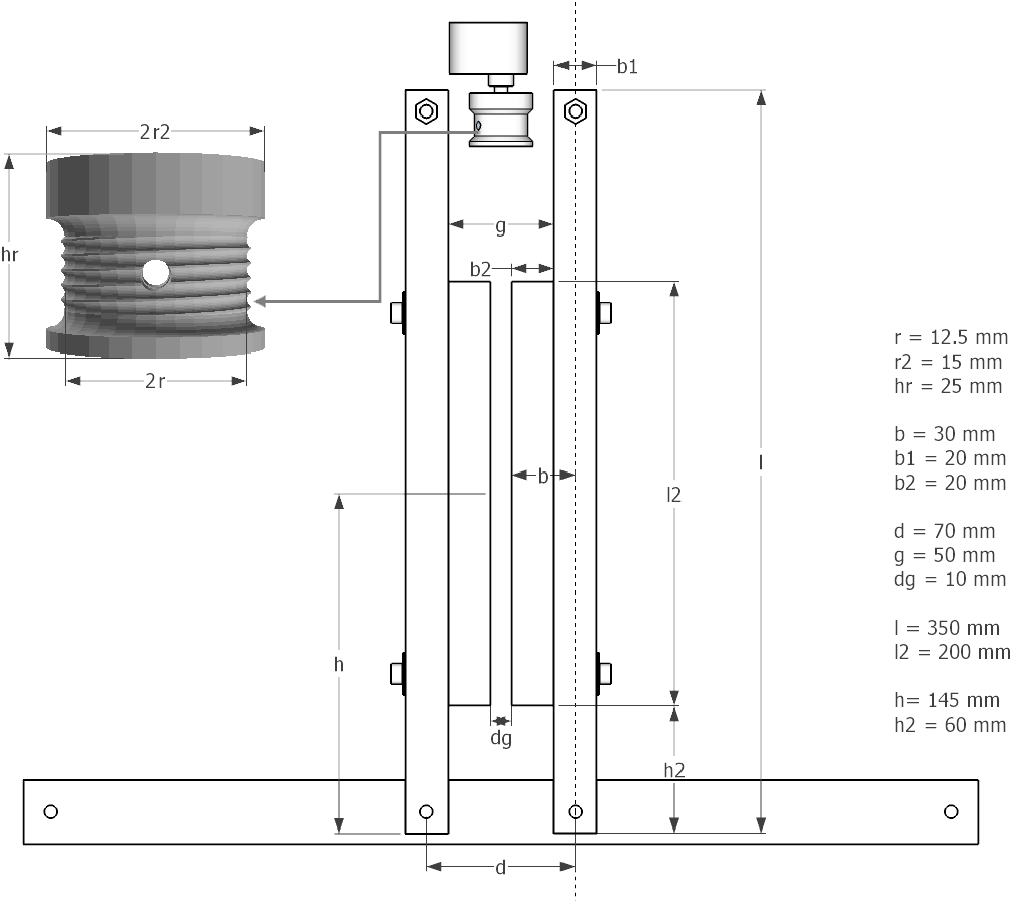
Schematic of the eARM system with arms in the maximum compression position. The design will require adjustments.

**Figure 16.**
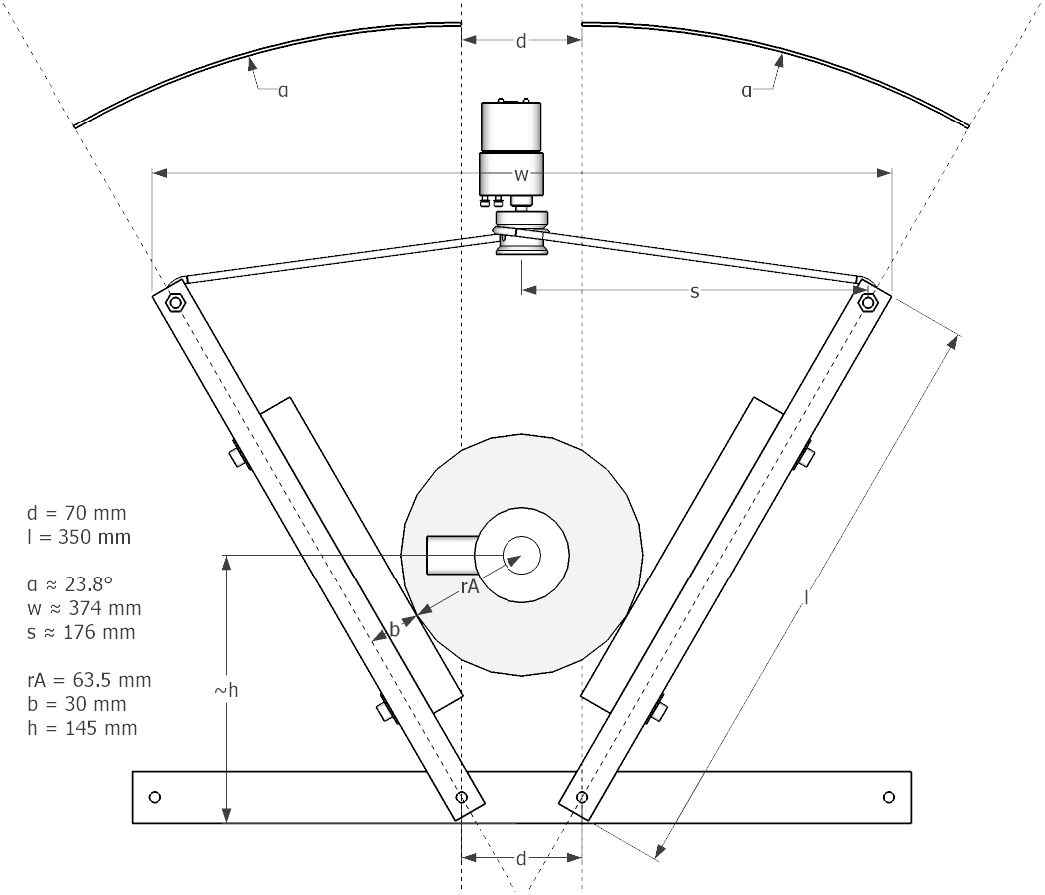
Schematic of the eARM system with minimum compresion showing the bag-valve resuscitator. The design will require adjustments.

**Figure 17.**
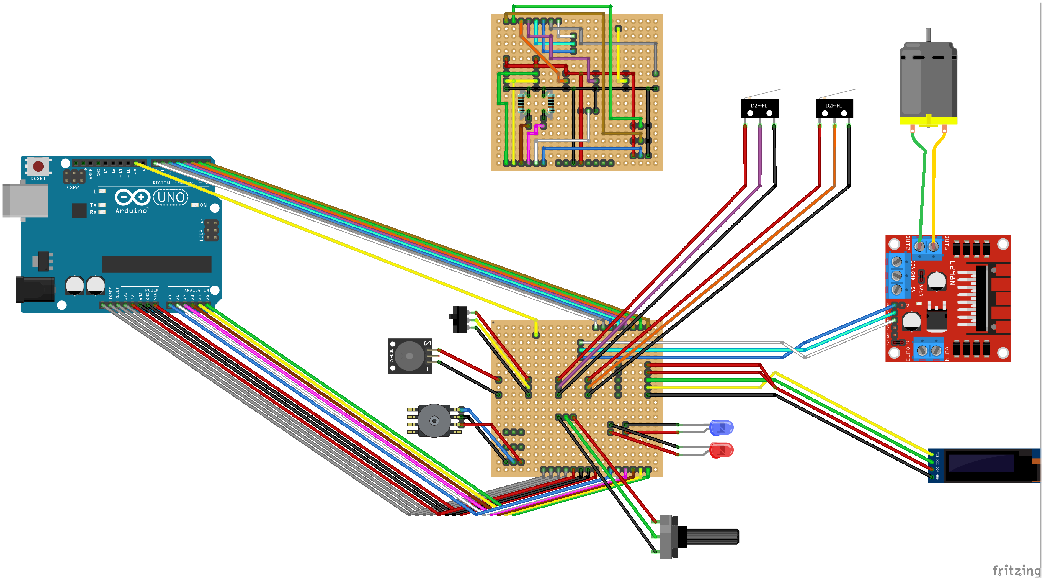
Schematic of the electrical connections for eARM system. The design will require adjustments.

## Data Availability

All the data to produce the plots in the manuscript can be requested from

## Acknowledgements

We acknowledge financial support from the Minister for Higher Education, Research and the Arts of the German State of Hessen and from the Sparkasse Marburg-Biedenkopf. ECC wants to thank financial support through an Experienced Reseach Fellowship from the Alexander von Humboldt Foundation.

## Author contributions statement

ECC proposed the study supported by MK. MK coordinated the workgroup. ECC proposed the CLAPER design with input from GGHC, BS and ARM. CM and JO proposed the eARM design, which was further developed, built and programmed jointly by CM, JO, JT and JW. Programming and documentation of eARM was supported also by SK, CS and DG. The designs were refined with medical input from TW and BB, as well as technical input from KK and GK. The systems were characterized by ECC, GGHC, BS, JO, CM and KK. FH and JN provided support to aspects as information recopilation, distribution and coordination between the medical, mechanical/software developers and test experts. The manuscript was written by ECC, JO, CM and JN. All authors reviewed the manuscript.

